# An in-depth investigation of the safety and immunogenicity of an inactivated SARS-CoV-2 vaccine

**DOI:** 10.1101/2020.09.27.20189548

**Authors:** Jing Pu, Qin Yu, Zhifang Yin, Ying Zhang, Xueqi Li, Dandan Li, Hongbo Chen, Runxiang Long, Zhimei Zhao, Tangwei Mou, Heng Zhao, Shiyin Feng, Zhongping Xie, Lichun Wang, Zhanlong He, Yun Liao, Shengtao Fan, Qiongzhou Yin, Ruiju Jiang, Jianfeng Wang, Lingli Zhang, Jing Li, Huiwen Zheng, Pingfang Cui, Guorun Jiang, Lei Guo, Mingjue Xu, Huijuan Yang, Shan Lu, Xuanyi Wang, Yang Gao, Xingli Xu, Linrui Cai, Jian Zhou, Li Yu, Zhuo Chen, Chao Hong, Dan Du, Hongling Zhao, Yan Li, Kaili Ma, Yunfei Ma, Donglan Liu, Shibao Yao, Changgui Li, Yanchun Che, Longding Liu, Qihan Li

**Author notes:** **Co-first authors:** Jing Pu, Qin Yu, Zhifang Yin, Ying Zhang, Xueqi Li, Dandan Li and Hongbo Chen contributed equally to this study. **Corresponding author:** Qihan Li, MD, PhD, Institute of Medical Biology, Chinese Academy of Medical Sciences and Peking Union Medical College. 935 Jiaoling Road, Kunming, Yunnan, China, 650118., Longding Liu, PhD, Institute of Medical Biology, Chinese Academy of Medical Sciences and Peking Union Medical, College., Yanchun Che, MS, Institute of Medical Biology, Chinese Academy of Medical Sciences and Peking Union Medical College., Changgui Li, PhD, National Institute of Food and Drug Control, China.

## Abstract

**BACKGROUND:** In-depth investigations of the safety and immunogenicity of inactivated SARS-CoV-2 vaccines are needed.

**METHOD:** In a phase I randomized, double-blinded, and placebo-controlled trial involving 192 healthy adults 18-59 years of age, two injections of three different doses (50 EU, 100 EU and 150 EU) of an inactivated SARS-CoV-2 vaccine or the placebo were administered intramuscularly with a 2- or 4-week interval between the injections. The safety and immunogenicity of the vaccine were evaluated within 28 days.

**FINDING:** In this study, 191 subjects assigned to three doses groups or the placebo group completed the 28-day trial. There were 44 adverse reactions within the 28 days, most commonly mild pain and redness at the injection site or slight fatigue, and no abnormal variations were observed in 48 cytokines in the serum samples of immunized subjects. The serum samples diluted from 1:32 to 1:4096 and incubated with the virus did not show antibody-dependent enhancement effects (ADEs) with regard to human natural killer cells, macrophages or dendritic cells. At day 14, the seroconversion rates had reached 92%, 100% and 96% with geometric mean titers (GMTs) of 18.0, 54.5 and 37.1, and at day 28, the seroconversion rates had reached 80%, 96% and 92% with GMTs of 10.6, 15.4 and 19.6in 0, 14 and 0, 28 procedures, respectively. Seroconversion was associated with the synchronous upregulation of ELISA antibodies against the S protein, N protein and virion and a cytotoxic T lymphocyte (CTL) response. Transcriptome analysis shaped the genetic diversity of immune response induced by the vaccine.

**INTERPRETATION:** In a population aged 18-59 years, this inactivated SARS-CoV-2 vaccine was safe and immunogenic.

**Trial registration:** NCT04412538

**FUNDING:** The National Key R&D Program of China (2020YFC0849700), the Program of Chinese Academy of Medicine Science and the Major Science and Technology Special Projects of Yunnan Province.

---

Coronavirus disease 2019 (COVID-19) is caused by a novel member of the Coronaviridae family called severe acute respiratory syndrome coronavirus 2 (SARS-CoV-2). From the emergence of COVID-19 at the end of 2019 to September 2020, more than 26 million cases and more than 800,000 deaths had been recorded, indicating that COVID-19 poses a substantial threat to public health worldwide ^1^. Because of the highly contagious nature of SARS-CoV-2 and the severe clinical outcomes ^2,3^, one of the primary strategies to control the pandemic is to develop an effective vaccine, and within a short period, clinical trials of several vaccines have been initiated ^4-7^. To date, the data obtained from phase I/II clinical trials have focused on serological detection to assess the immunogenicity of these vaccines ^4,7,8^. The data suggest that SARS-CoV-2, an enveloped virus, possesses various encoded antigenic components, including S (spike), N (nucleocapsid), E (envelope) and M (membrane) antigens ^9^, all of which might theoretically be recognized by the immune system during infection; however, the key antigen is the S protein, which mediates virion entry into cells by interacting with the ACE2 receptor ^10^. Our previous work, based on an analysis of the composition of antibodies in convalescent serum from COVID-19 patients, suggested a technical strategy for the preparation of an inactivated SARS-CoV-2 vaccine in which the inactivation process yields an inactivated virion exposing the S, N and other viral antigens. This vaccine was found to effectively elicit immune protection in nonhuman primates under viral challenge and was approved for a phase I clinical trial (permission number: 2020L00020 by the Chinese Food and Drug Administration (CFDA); clinical trial registration number: NCT04412538). Here, we further investigated the safety and immunogenicity of this vaccine in immunized individuals in a phase I trial, especially focusing on safety with regard to the immunopathology of the vaccine. The results obtained are encouraging, and further study is needed.

## Materials and Methods

### Viruses and cells

All SARS-CoV-2 virus strains used in this work were isolated from hospitalized patients including indigenous and imported cases confirmed COVID-19 in Yunnan Hospital of Infectious Diseases from January to May 2020. The strain with mutation of D614G in S protein was isolated from a child patient who returned from U.S. to hometown and was identified being infected by SARS-CoV-2 through clinical diagnostic and q-RT-PCR in March, 2020. The virus was proliferated in Vero cells (WHO) and was titrated with a microtitration assay. Vero cells were cultured in Dulbecco’s modified Eagle’s medium (DMEM; Corning, NY, USA) containing 5% fetal bovine serum (FCS; HyClone, Logan, USA).

### Inactivated vaccine

The SARS-CoV-2 inactivated vaccine was developed by the Institute of Medical Biology (IMB), Chinese Academy of Medical Sciences (CAMS). Briefly, the virus strain, named KMS-1 (GenBank No: MT226610.1), was inoculated into Vero cells for production in an environment that met the BSL requirements. The harvested viruses were inactivated by formaldehyde (v:v=1:4000) for 48 h to lyse the viral membrane, purified via chromatography and concentrated. A second inactivation with beta-propiolactone (v:v=1:2000) was performed to destroy the viral genomic structure, followed by a second purification and concentration using the same protocol. The vaccine stock was evaluated using various quality indexes, including antigen content, immunogenicity, sterility and residue testing. The viral antigen content was measured via ELISA. The vaccine contained 50, 100 or 150 EU of inactivated viral antigen adsorbed to 0.25 mg of Al(OH)_3_ adjuvant and suspended in 0.5 ml of buffered saline for each dose. The placebo contained only the same amount of Al(OH)_3_ in buffer.

### Study design and participants

The trial was designed based upon the principles of randomization, double-blinding and placebo control. The study protocol was reviewed and approved by the Ethics Committee of the West China Second University Hospital, Sichuan University. An independent data safety monitoring board was established to oversee the safety data during the study, specifically during the 7 days after each inoculation (p.i.). The trial was conducted according to the principles of the Declaration of Helsinki and Good Clinical Practice at the West China Second University Hospital, Sichuan University. Healthy volunteers 18 to 59 years of age were eligible for enrollment after providing informed consent. The inclusion and exclusion criteria are listed in the supplementary appendix. The enrolled participants were randomly assigned at a ratio of 1:1 to receive two inoculations at an interval of 14 days or 28 days, and the subjects in each schedule were assigned at a ratio of 1:1:1:1 to receive one of the three vaccine doses (50 EU, 100 EU and 150 EU) or the placebo. All the enrolled participants were asked to record solicited and unsolicited adverse events, if any, for a period of 28 days. Study staff visited participants on site to track their health status and determine whether they needed medical care. Blood samples were taken from the enrolled participants at days 0 (baseline), 7, 14 and 28 (0, 14 schedule) or 7 and 28 (0, 28 schedule) days postimmunization to evaluate the immunogenicity of the vaccine at different time points and to determine the vaccine safety profile through the detection of 48 cytokines in the serum, the assessment of possible ADEs with regard to viral infection and the analysis of mRNA gene expression in peripheral blood mononuclear cells (PBMCs).

### End points of the clinical trial

The primary end point was the total rate of adverse reactions from 0 to 28 days postimmunization. The secondary end points were serological evidence of the immunogenicity of the vaccine.

### Statistical analysis

Descriptive statistics (means and standard deviations for normally distributed variables, medians and IQRs for nonnormally distributed variable) were used for continuous variables, while frequencies and proportions were used for categorical variables. The safety analysis was performed with the data from the participants who had received at least one dose of the vaccine and for whom safety data were available. The numbers and proportions of participants with adverse reactions or events were summarized. The immunogenicity analysis was conducted with the data from the full cohort, including all participants who received injections and had results for the antibody test. The antibodies against SARS-CoV-2 are summarized as geometric mean titers with 95% CIs, and the cellular responses are presented as the proportion of positive responders. The chi-square test or Fisher’s exact test was used to analyze the categorical data, and the t-test or Wilcoxon rank-sum test (for nonnormally distributed data) was used to compare log-transformed antibody titers. A P value lower than 0.05 (two-sided) was considered to be significant. The statistical analysis was performed by an independent statistician using GraphPad Prism software (San Diego, CA, USA) and STATA software (Version 15.0; STATA Corp., College Station, TX, USA).

### Laboratory detection

#### Neutralizing antibody test

The neutralizing antibody assay was performed via microtitration, and the neutralizing titer in the sera was determined by CPE observation. A brief description is attached in the supplemental appendix.

#### ELISA

ELISAs were conducted with antibodies against the S protein, N protein and virions that were developed by this institute. A brief description can be found in the supplemental appendix.

#### ELISpot

An ELISpot assay was performed with a Human IFN-γ ELISpot Kit (Mabtech, Cincinnati, OH, USA) according to the manufacturer’s protocol. A brief description is attached in the supplemental appendix.

#### Immune cell populations

The isolation of immune cell populations was performed according to a standard protocol. A brief description can be found in the supplemental appendix.

#### Cytokine assay

Detection of 48 cytokines in human peripheral blood was performed according to the kit manufacturer’s instructions. A brief description is attached in the supplemental appendix.

#### Transcriptome assay

Transcriptome assays were performed by Novogene Co., Ltd., China. To exclude individual differences, each group included two samples (A and B) at each time point, and each sample was mixed with PBMCs from five individuals. The raw microarray data were submitted to the National Genomics Data Center (NGDC) and are available (PRJCA003531). A brief description is attached in the supplemental appendix.

#### ADE detection

Human NK cells, DCs, and mononuclear/macrophages isolated from donors who provided their informed consent were infected by the virus in different multiplicities of infection (MOIs) in the presence of mixed sera from 12 immunized individuals that had the same neutralizing titer and were diluted from 1:32 to 1:4096. Antibody-dependent enhancement (ADE) was determined via viral load detection by q-RT-PCR at various time points. A brief description is attached in the supplemental appendix.

#### qRT-PCR

The RNA extraction and qRT-PCR assay was according standard protocol. The brief description can be seen in supplemental appendix.

## Results

### Study participants

A total of 294 adults aged 18 to 59 years were evaluated for inclusion in this phase I trial. Among them, 102 persons were excluded, of whom 84 were ineligible, and 18 withdrew their informed consent. The remaining 192 participants were randomly assigned at a ratio of 1:1 to receive two inoculations with an intervening interval of 14 days or 28 days, and subjects for each schedule were assigned at a ratio of 1:1:1:1 to receive one of the three doses of the vaccine or the placebo (Fig. 1).

**Fig. 1.**
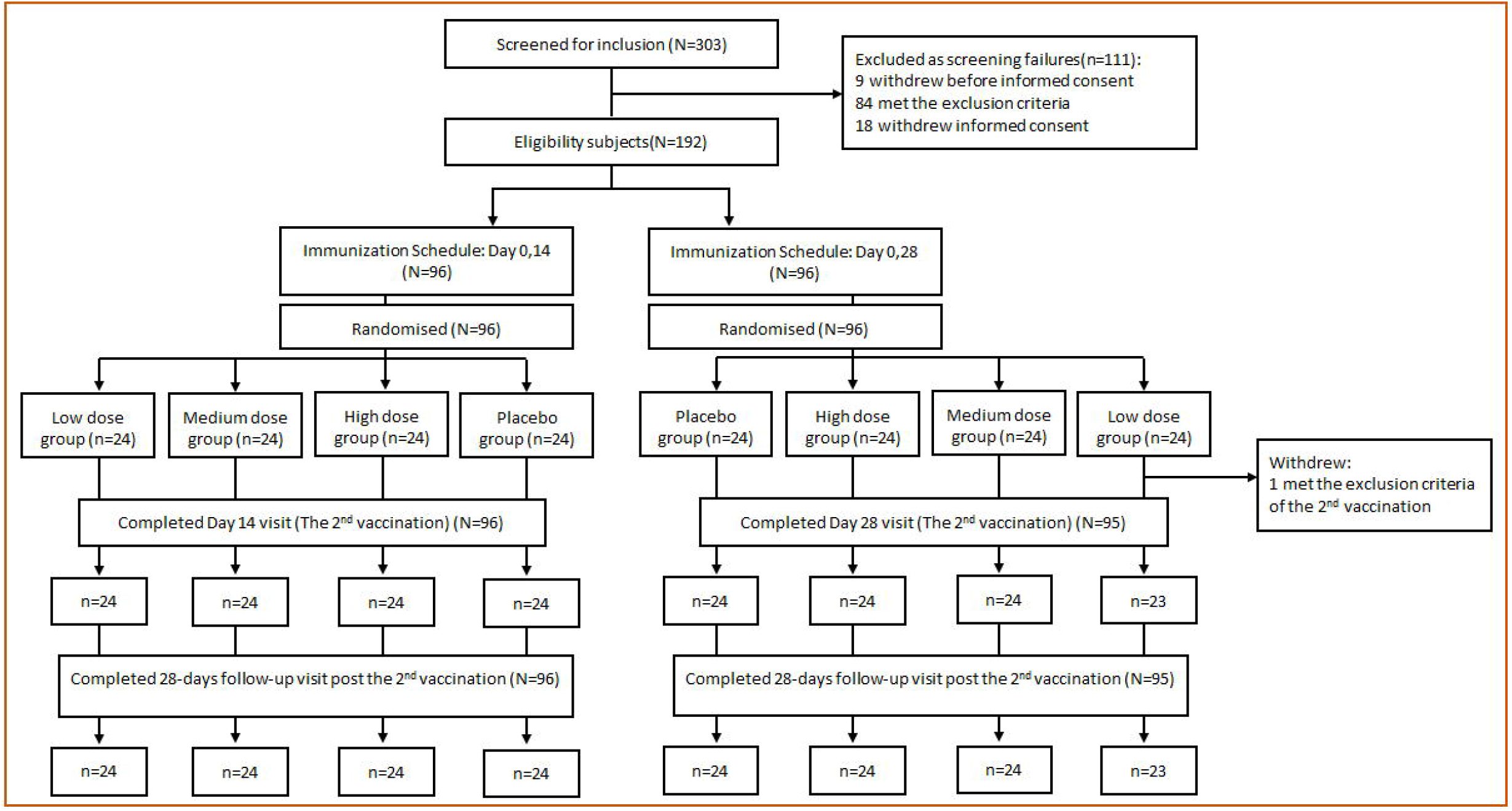
Screening, randomization and inclusion in phase I clinical trial.

From May 2020 through August 2020, all participants who received two inoculations of the vaccine or placebo were monitored for any clinical manifestations and were required to provide blood samples 3 (0, 14 schedule) or 2 (0, 28 schedule) times after inoculation. The withdrawal rate was 0.5%: 1 participant in the low-dose group who was assigned to the 0, 28 schedule did not receive the second dose. The 191 participants were divided as follows: 24 in each of the three different dose groups and the control group assigned to the 0, 14 schedule and 23, 24, 24 and 24 in the low-dose, medium-dose, high-dose and control groups assigned to the 0, 28 schedule (Fig. 1). The demographic characteristics of the participants in each group are shown in Table 1.

**Table 1.**
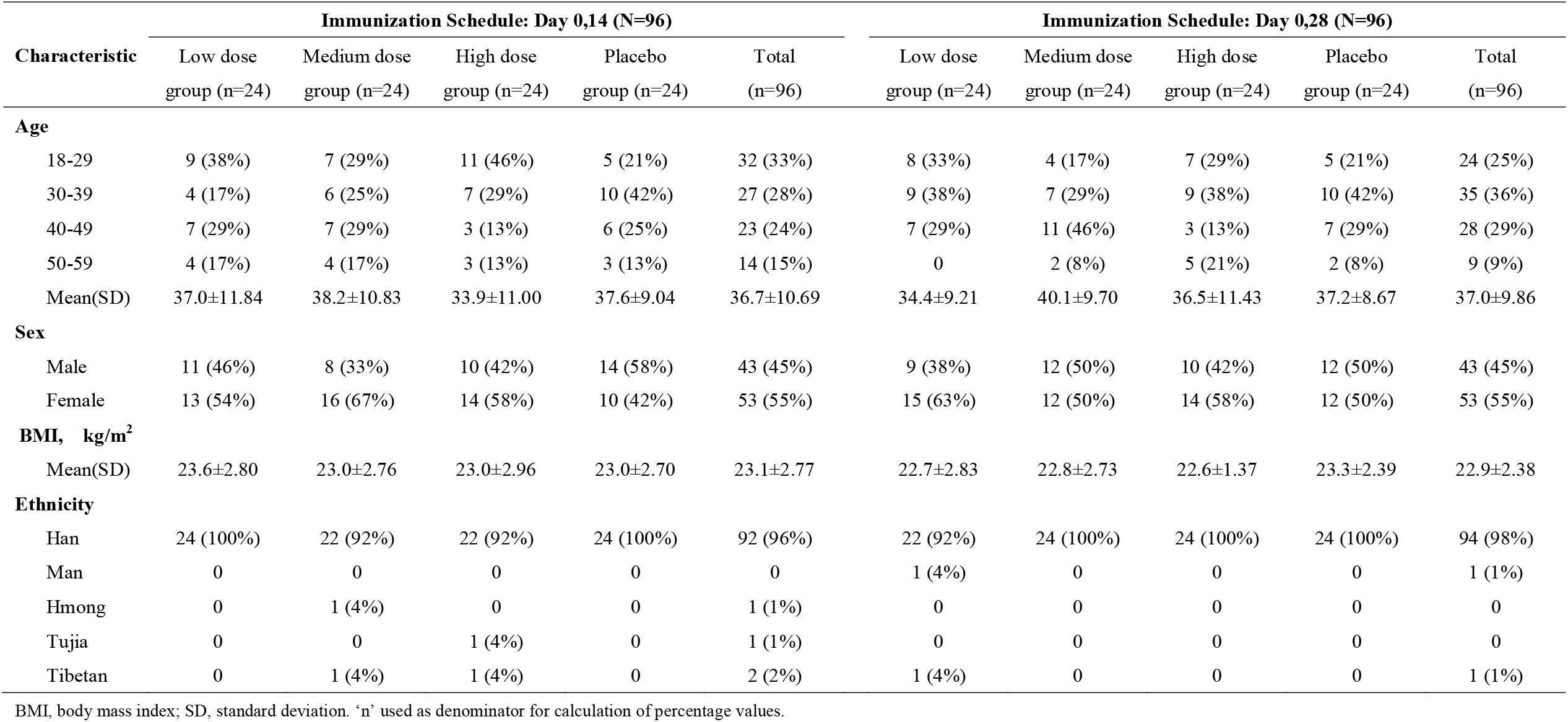
Baseline characteristics of the study participants.

| Characteristic | Immunization Schedule: Day 0,14 (N=96) |  |  |  |  | Immunization Schedule: Day 0,28 (N=96) |  |  |  |  |
| --- | --- | --- | --- | --- | --- | --- | --- | --- | --- | --- |
|  | Low dose<br>group (n=24) | Medium dose<br>group (n=24) | High dose<br>group (n=24) | Placebo<br>group (n=24) | Total<br>(n=96) | Low dose<br>group (n=24) | Medium dose<br>group (n=24) | High dose<br>group (n=24) | Placebo<br>group (n=24) | Total<br>(n=96) |
| **Age** |  |  |  |  |  |  |  |  |  |  |
| 18-29 | 9 (38%) | 7 (29%) | 11 (46%) | 5 (21%) | 32 (33%) | 8 (33%) | 4 (17%) | 7 (29%) | 5 (21%) | 24 (25%) |
| 30-39 | 4 (17%) | 6 (25%) | 7 (29%) | 10 (42%) | 27 (28%) | 9 (38%) | 7 (29%) | 9 (38%) | 10 (42%) | 35 (36%) |
| 40-49 | 7 (29%) | 7 (29%) | 3 (13%) | 6 (25%) | 23 (24%) | 7 (29%) | 11 (46%) | 3 (13%) | 7 (29%) | 28 (29%) |
| 50-59 | 4 (17%) | 4 (17%) | 3 (13%) | 3 (13%) | 14 (15%) | 0 | 2 (8%) | 5 (21%) | 2 (8%) | 9 (9%) |
| Mean(SD) | 37.0±11.84 | 38.2±10.83 | 33.9±11.00 | 37.6±9.04 | 36.7±10.69 | 34.4±9.21 | 40.1±9.70 | 36.5±11.43 | 37.2±8.67 | 37.0±9.86 |
| **Sex** |  |  |  |  |  |  |  |  |  |  |
| Male | 11 (46%) | 8 (33%) | 10 (42%) | 14 (58%) | 43 (45%) | 9 (38%) | 12 (50%) | 10 (42%) | 12 (50%) | 43 (45%) |
| Female | 13 (54%) | 16 (67%) | 14 (58%) | 10 (42%) | 53 (55%) | 15 (63%) | 12 (50%) | 14 (58%) | 12 (50%) | 53 (55%) |
| **BMI, kg/m<sup>2</sup>** |  |  |  |  |  |  |  |  |  |  |
| Mean(SD) | 23.6±2.80 | 23.0±2.76 | 23.0±2.96 | 23.0±2.70 | 23.1±2.77 | 22.7±2.83 | 22.8±2.73 | 22.6±1.37 | 23.3±2.39 | 22.9±2.38 |
| **Ethnicity** |  |  |  |  |  |  |  |  |  |  |
| Han | 24 (100%) | 22 (92%) | 22 (92%) | 24 (100%) | 92 (96%) | 22 (92%) | 24 (100%) | 24 (100%) | 24 (100%) | 94 (98%) |
| Man | 0 | 0 | 0 | 0 | 0 | 1 (4%) | 0 | 0 | 0 | 1 (1%) |
| Hmong | 0 | 1 (4%) | 0 | 0 | 1 (1%) | 0 | 0 | 0 | 0 | 0 |
| Tujia | 0 | 0 | 1 (4%) | 0 | 1 (1%) | 0 | 0 | 0 | 0 | 0 |
| Tibetan | 0 | 1 (4%) | 1 (4%) | 0 | 2 (2%) | 1 (4%) | 0 | 0 | 0 | 1 (1%) |

BMI, body mass index; SD, standard deviation. 'n' used as denominator for calculation of percentage values.

### Comprehensive observation of adverse reactions

Because knowledge of the pathogenesis of SARS-CoV-2 infection is still lacking ^11,12^, in this trial, the specific vaccine safety concern was abnormal immunopathologic events. We observed not only solicited clinical adverse reactions for 7 days after each inoculation and unsolicited events for 28 days after the full vaccination but also the variations in cytokine levels in the serum of 50% of the subjects inoculated with the vaccine or placebo. Furthermore, based on the serious concern regarding a possible ADE effect in vaccine development ^13^, the immune serum samples from vaccinated individuals were investigated across a range of dilutions for possible ADE. The 18 solicited systemic adverse reactions observed in the 7 days after each inoculation were distributed as follows: 3, 1, 1 and 0 in the low-dose, medium-dose, high-dose and placebo groups assigned to the 0 and 14 schedule and 2, 4, 4, and 3 in those groups assigned to the 0 28 schedule. Unsolicited reactions were reported by 8.3%, 0.0%, and 8.3% of the participants in the low-, medium- and high-dose groups compared to 0.0% in the placebo group among those assigned to the 0, 14 schedule, while they were reported by 0.0%, 8.3%, and 12.5% of the participants in the low-, medium- and high-dose groups compared to 4.2% in the placebo group among those assigned to the 0, 28 schedule. The most common adverse reactions were mild pain and redness at the injection site and slight fatigue. No severe (grade 3) adverse reactions or serious reactions were observed within 28 days (Table 2). Furthermore, the levels of various cytokines in the sera of immunized subjects in the 3 dose groups following the 2 schedules did not show abnormalities compared to those in the sera from subjects in the placebo groups (Fig. 2). There were no significant differences between the vaccine and placebo groups with regard to the counts of various T cell populations in the peripheral blood (Supplemental Fig. 1). To investigate possible ADE in individuals immunized with the inactivated vaccine, immune sera collected from 15 subjects with similar neutralizing antibody titers were mixed and diluted from 1:32 to 1:4096 and then incubated with SARS-CoV-2 for inoculating the cultured human DCs, NK cells and mononuclear cells/macrophages followed by the detection of dynamic viral proliferation in the cells and supernatant. The results did not show variations of dynamic viral proliferation with existence of immune sera rather than 1-2 fold compared to viral control, and suggest that the immunized sera in the dilution range of 1:32 to 1:4096 do not lead to ADE in these human immune cells compared with sera from individuals in the placebo group and the positive Dengue virus control group (Fig. 3). These results suggest that there are no immunopathologic events related to vaccination.

**Table 2.** Adverse events within 28 days after vaccination in the safety population.

| Adverse reactions | 0,14 procedure |  |  |  | 0,28 procedure |  |  |  |
| --- | --- | --- | --- | --- | --- | --- | --- | --- |
|  | Low dose | Middle dose | High dose | Placebo | Low dose | Middle dose | High dose | Placebo |
|  | group (n=24) | group (n=24) | group (n=24) | group (n=24) | group (n=24) | group(n=24) | group(n=24) | group(n=24) |
|  | *number of participants (%)* |  |  |  | *number of participants (%)* |  |  |  |
| All adverse reactions within 0-7 days | 8 (33.3) | 4 (16.7) | 4 (16.7) | 3 (12.5) | 5 (20.8) | 7 (29.2) | 9 (37.5) | 4 (16.7) |
| Solicited injection site adverse reactions within 0-7 days |  |  |  |  |  |  |  |  |
| Total | 7 (29.2) | 4 (16.7) | 2 (8.3) | 3 (12.5) | 3 (12.5) | 4 (16.7) | 5 (20.8) | 0 (0.0) |
| Pain | 6 (25.0) | 4 (16.7) | 2 (8.3) | 2 (8.3) | 3 (12.5) | 3 (12.5) | 5 (20.8) | 0 (0.0) |
| Itch | 2 (8.3) | 1 (4.2) | 0 (0.0) | 1 (4.2) | 0 (0.0) | 0 (0.0) | 0 (0.0) | 0 (0.0) |
| Redness | 1 (4.2) | 0 (0.0) | 0 (0.0) | 1 (4.2) | 0 (0.0) | 0 (0.0) | 0 (0.0) | 0 (0.0) |
| Swelling | 0 (0.0) | 0 (0.0) | 0 (0.0) | 0 (0.0) | 0 (0.0) | 1 (4.2) | 0 (0.0) | 0 (0.0) |
| Solicited systemic adverse reactions within 0-7 days |  |  |  |  |  |  |  |  |
| Total | 3 (12.5) | 1 (4.2) | 1 (4.2) | 0 (0.0) | 2 (8.3) | 4 (16.7) | 4 (16.7) | 3 (12.5) |
| Fatigue | 2 (8.3) | 1 (4.2) | 0 (0.0) | 0 (0.0) | 2 (8.3) | 3 (12.5) | 3 (12.5) | 2 (8.3) |
| Diarrhoea | 0 (0.0) | 1 (4.2) | 1 (4.2) | 0 (0.0) | 0 (0.0) | 0 (0.0) | 1 (4.2) | 0 (0.0) |
| Fever | 1 (4.2) | 0 (0.0) | 0 (0.0) | 0 (0.0) | 0 (0.0) | 1 (4.2) | 0 (0.0) | 0 (0.0) |
| Rash | 1 (4.2) | 0 (0.0) | 0 (0.0) | 0 (0.0) | 0 (0.0) | 0 (0.0) | 0 (0.0) | 1 (4.2) |
| Unsolicited adverse reactions within 0-7 days | 2 (8.3) | 0 (0.0) | 2 (8.3) | 0 (0.0) | 0 (0.0) | 2 (8.3) | 3 (12.5) | 1 (4.2) |
| Overall adverse reactions within 0-28 days | 8 (33.3) | 4 (16.7) | 4 (16.7) | 3 (12.5) | 6 (25.0) | 7 (29.2) | 9 (37.5) | 3 (12.5) |

No severe (grade 3) adverse reaction and serious adverse events were observed within 28 days

**Fig. 2.**
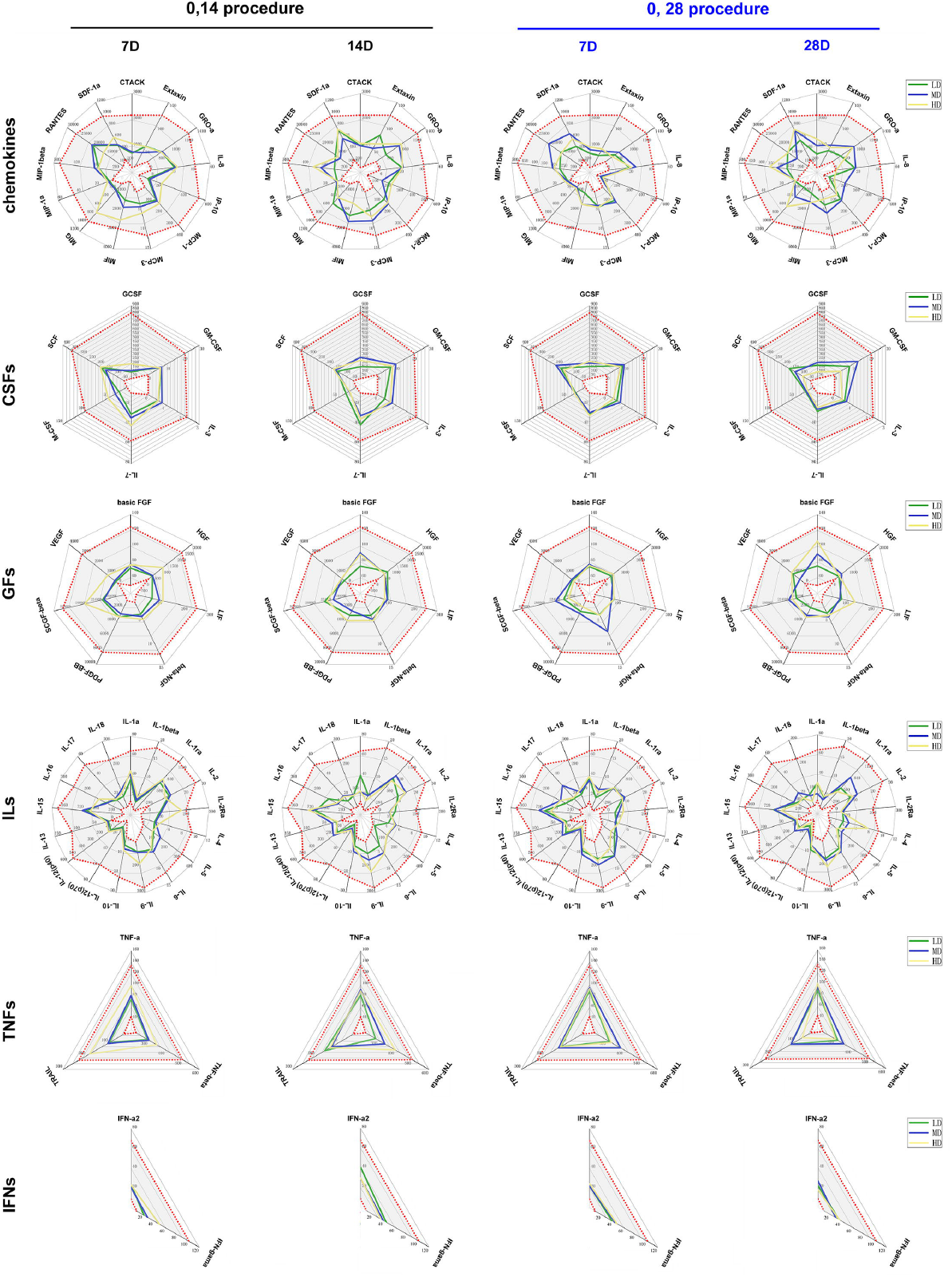
Variations in 48 cytokines in the serum of immunized individuals and the observation of ADE. Levels of 48 cytokines were monitored in the serum of the subjects who received the vaccine and placebo who were assigned to the 0, 14 schedule (black) or the 0, 28 schedule (blue). Cytokines were Chemokines, interleukins (ILs), growth factors (GFs), colony stimulating factors (CSFs), tumor necrosis factors (TNFs), interferon (IFNs). The levels of 48 cytokines (pg/mL) in the serum of subjects before receiving vaccine and placebo are shown as gray intervals between red spots in each figure. Control (Con, 0 EU), low dose (LD, 50 EU), middle dose (MD, 100 EU) and high dose (HD, 150 EU).

**Fig. 3.**
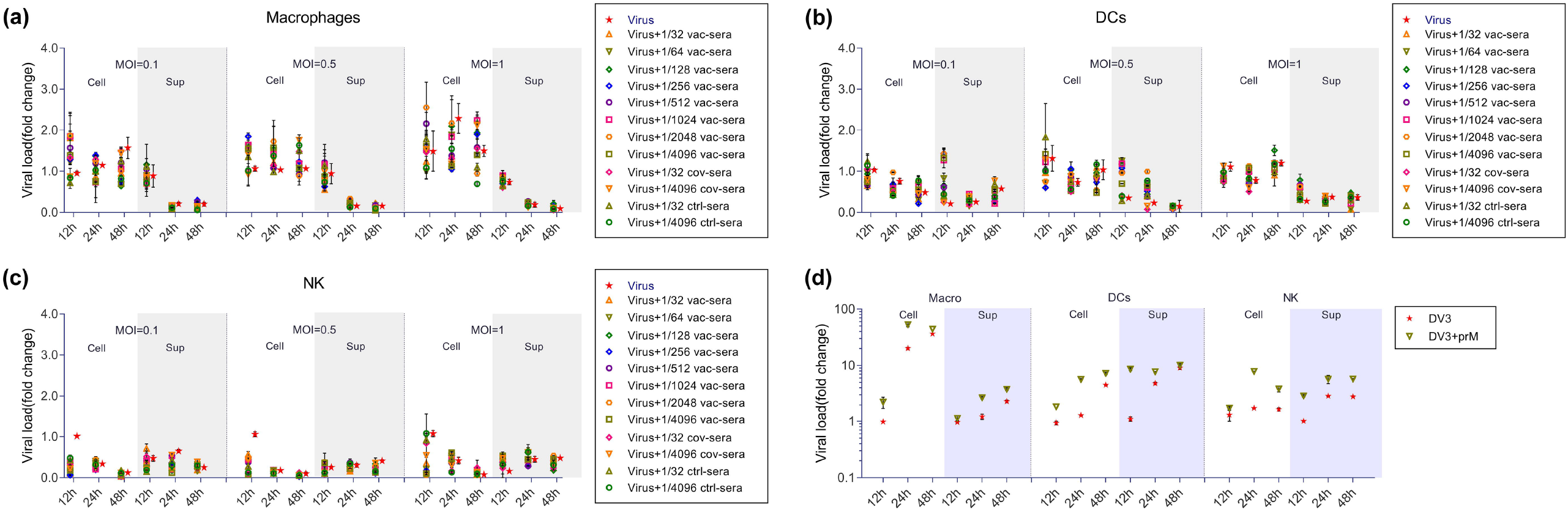
The observation of ADE. a-c: Fold change in relative viral load of supernatant and cell lysate of mononuclear cells/macrophages (a), DCs (b) and NK cells (c) from human PBMCs infected with SARS-CoV-2 at three MOI in the presence of a series of dilutions of vaccine immune serum (1:32, 1:64, 1:128, to 1:4096) quantified by real-time PCR. Viral load of cell lysate were normalized to viral copies in cell lysate of Virus group at 12 hours, and viral load of supernatant were normalized to viral copies in virus input at 0 hour. Vac-sera, vaccine immune antisera from human; cov-sera, convalescent antisera from COVID-19 patients; ctrl-sera, control sera from healthy human. Data are shown as the mean ± SEM. d: Fold change in relative viral load of supernatant and cell lysate of mononuclear cells/macrophages, DCs and NK cells from human PBMCs infected with DENV-3 in the presence of mAb against prM quantified by real-time PCR. Viral load of cell lysate were normalized to viral copies in cell lysate of Virus group at 12 hours, and viral load of supernatant were normalized to viral copies in virus input at 0 hour. Data are shown as the mean ± SEM.

### Assessment of immunity elicited in the 3 dose groups assigned to the 2 schedules

The results obtained for the three dose groups assigned to the two schedules with regard to the neutralizing antibody titers of the immunized subjects in the groups assigned to the 0, 14 schedule showed seroconversion rates of 55%, 100% and 88% in the low-, medium- and high-dose groups compared to the placebo group at day 7 after the booster, and their geometric mean titers (GMTs) increased by 5.0, 42.5 and 24.4, respectively. Furthermore, at day 14, the seroconversion rates in the three groups reached 92%, 100% and 96%, with GMTs of 18.0, 54.5 and 37.1, respectively (Fig. 4a). However, there appeared to be a decreasing trend in the neutralizing antibody titers from day 14 to day 28 (Fig. 4a). In the groups assigned to the 0, 28 schedule, the seroconversion rate of the neutralizing antibodies reached 80%, 96% and 92%, with GMTs of 10.6, 15.4 and 19.6, at day 28 after immunization, with an increasing trend starting from day 7 after immunization (Fig. 4b). Significantly, ELISA with antibodies against the S protein, N protein and virion showed essentially synchronous increases regardless of the schedule to which the participants were assigned (Fig. 4a, b). The specific positive CTL responses against the S protein, N protein and virion in the ELISpot assay indicated a distinct increase at day 28 after the booster for both schedules (Fig. 4a, b). These results suggest that the vaccine elicits a synchronous dynamic response involving antibodies and CTLs against the viral antigens. The subsequent serological detection in the available serum samples of 23 subjects immunized with the medium dose of the vaccine at 90 days after the second inoculation suggested that the seroconversion rate of the neutralizing antibodies was 91.3% with a GMT of 8.5, while the ELISA antibodies showed 100% of seroversion including anti-S, anti-N and anti-virion antibodies of 91%, 61% and 83% with GMTs of 656, 594 and 691, respectively (Fig. 4c). The IgG1 subtype was the common subtype against all three antigens (Table S1). Additionally, the immune sera were found to neutralize the pandemic strains in North America (Fig. 4d) with the D614G mutation in the S protein (Supplemental Fig. 2) ^14^. To characterize the immunogenicity of the vaccine, we performed a comparative analysis of the mRNA profile of PBMCs from immunized individuals of medium-dose group in two schedules at day 7 and 28 after full immunization to shape the genetic diversity of the immune response elicited by the vaccine. The results suggest that the vaccine initiates and promotes a series of transcriptional activities in immune cells, which lead to the significant upregulation of many genes related to the immune response. The classification of all the different upregulated genes into various immune functions suggests that the immunization by the vaccine of medium-dose in two schedules results in the specific activation of the innate and adaptive immune responses compared to the placebo control. However, the expression of certain significant cytokine genes, including IL-6, IL-1, IL-2, TNF-α and IFN-γ, which were found to be distinctly increased in the peripheral blood of COVID-19 patients, varied only slightly compared to those in the placebo controls (Fig. 5a). Furthermore, our data indicate that the genes related to T and B cell activation were upregulated by approximately 40% and 25%, respectively, at day 7 after the second inoculation and varied dynamically during the periods from day 7 to day 14 or from day 7 to day 28 in two schedules (Fig. 5b, c). The genes related to the activation of DCs, mononuclear cells/macrophages and NK cells were upregulated to varying degrees in the same periods (Fig. 5d, e, f). All these results describe a specific genetic background of the immune response elicited by the vaccine.

**Fig. 4.**
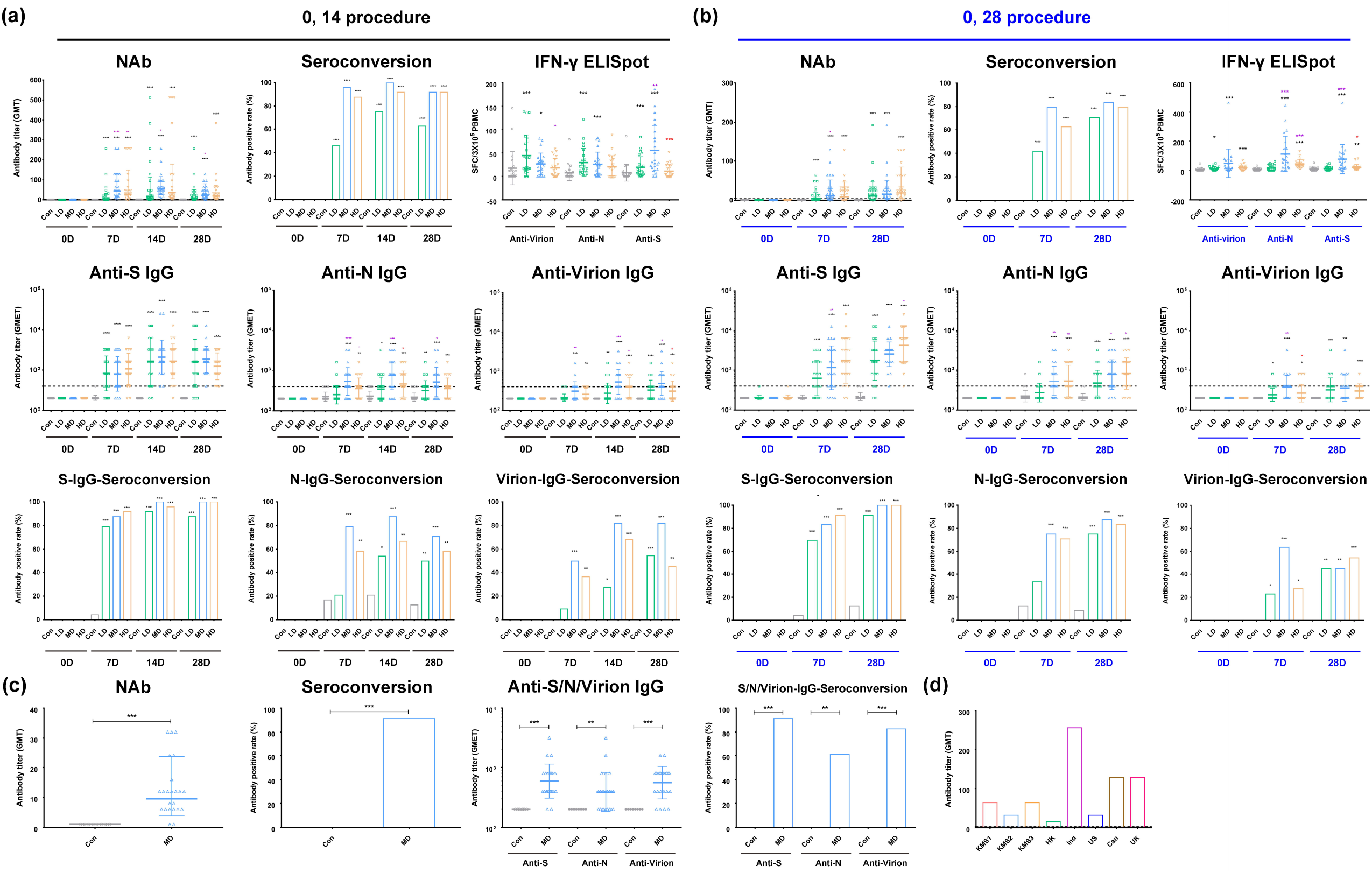
Immune response induced in immunized human individuals with inactivated SARS-Cov-2 vaccine in the 0, 14 and 0, 28 schedules. a. Neutralizing antibodies, ELISA antibodies (IgGs) against S protein, N protein and virion, and the specific positive CTL responses against the S, N and virion antigens induced by the inactivated vaccine in individuals assigned to the 0, 14 schedule after booster immunization. b. Neutralizing antibodies, ELISA antibodies (IgGs) against S protein, N protein and virion, and the specific positive CTL responses against the S, N and virion antigens induced by the inactivated vaccine in individuals assigned to the 0, 28 schedule after booster immunization. c. Neutralizing antibodies, ELISA antibodies (IgGs) against S protein, N protein and virion induced by the inactivated vaccine in individuals assigned to the 0, 28 schedule on 90 days after booster immunization. d. Neutralizing antibodies induced by the inactivated vaccine could identify pandemic strains from all over the world. Control (Con, 0 EU), low dose (LD, 50 EU), middle dose (MD, 100 EU) and high dose (HD, 150 EU). The antibody positive judgment threshold is marked with a dotted line in the figure. *, 0.01<p<0.05; **, 0.001<p<0.01; ***, p<0.001. The significant differences compared to the control group (Con) are shown in black, those compared to the low-dose group in purple, and those compared to the middle-dose group in red.

**Fig. 5.**
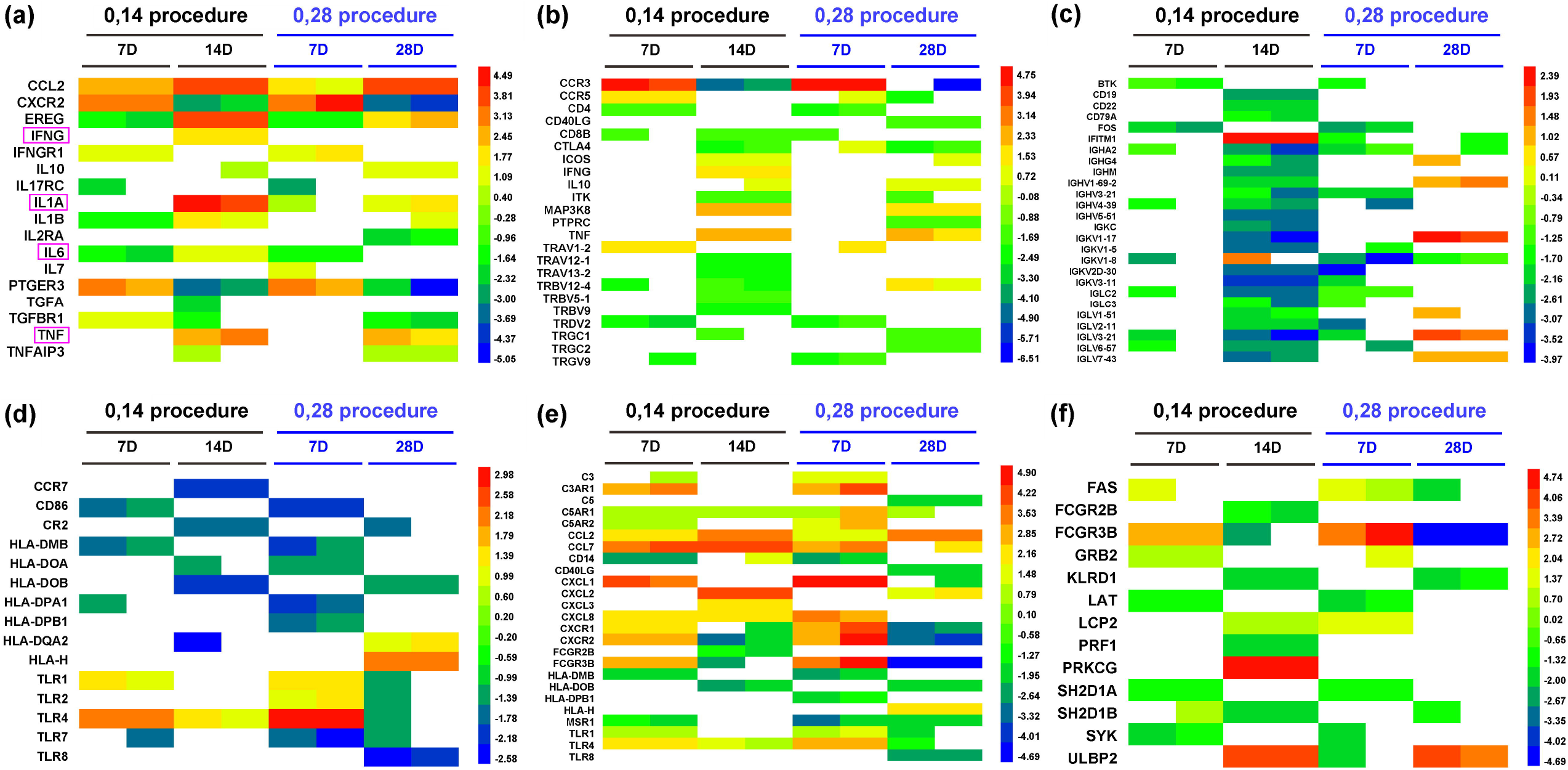
Genetic diversity of genes related to the immune response induced by the inactivated SARS-Cov-2 vaccine. a. The fold changes in some of the differentially expressed genes involved in cytokine production. Some important genes reported relating to COVID-19 were marked with the pink rectangle. b. The fold changes in some of the differentially expressed genes involved in T cell activation. c. The fold changes in some of the differentially expressed genes involved in B cell activation. d. The fold changes in some of the differentially expressed genes involved in DC cell activation. e. The fold changes in some of the differentially expressed genes involved in mononuclear cell/macrophage activation. f. The fold changes in some of the differentially expressed genes involved in NK cell activation. Each row represents one gene, and the samples are depicted in the columns. Red indicates genes that were expressed at higher levels, and blue denotes genes that were expressed at lower levels compared with the control group at the same time point. The color bars represent the log2 fold change.

## Discussion

Given the urgent need to control the COVID-19 pandemic, vaccine development is being accelerated into the clinical trials phase ^4,7,8^, even though understanding of the immunologic features of the antigens of SARS-CoV-2 remains poor. In this phase I trial, a study was performed to investigate the safety and immunogenicity of this inactivated vaccine in 191 subjects. The data collected show several notable features. First, the clinical safety observations among the 191 subjects suggest that there were no severe adverse reactions related to vaccination, and the most frequently reported events were mild, including redness, itching and swelling at the inoculation site and a few cases of slight fatigue; there were no significant differences between the vaccine and control groups. These data support the clinical safety of this vaccine. However, based on the current concern about the possibility of immunopathology due to SARS-Cov-2 infection ^13^, we extended our safety observation to the investigation of variations in cytokine levels in the serum, with particular attention paid to the possibility of ADE related to antibodies induced by vaccination ^12,15^. The test results suggested that there were no abnormalities in most of the 48 detected cytokines. In the dilution range of 1:32 to 1:4096, no ADE was observed in the virus-infected DCs, NK cells or mononuclear cells/macrophages in the immune sera from vaccinated individuals. Although we cannot conclude that this vaccine will not cause ADE, these observations at least suggest that the likelihood of ADE as a result of inoculation with this vaccine is small. Second, serological detection showed not only neutralizing antibody but also ELISA antibodies against the S protein, N protein and complete virion antigens elicited in the vaccinated population, and there were dynamic alterations based on the dose and vaccination schedule. However, the medium and high doses in both the 0, 14 and 0, 28 schedule groups led to 100% seroconversion of ELISA anti-S antibody after two inoculations, and interestingly, the medium dose group assigned to the 0, 14 schedule reached 100% seroconversion of the neutralizing antibody with the highest GMT value. Importantly, the neutralizing antibody can neutralize different pandemic strains with diverse mutations. At the time of the antibody response, a CTL response with IFN-r specificity against the S, N and virion antigens was detected in immunized individuals in comparison with individuals receiving placebo, which suggests that any one of three antigens enables the specific activation of T cells, even if they do not show dose-dependent effects. These immunological indexes indicate a systemic immune response elicited by our vaccine in the human population. To shape the genetic diversity of the specific immunity elicited by the vaccine, we examined the mRNA gene profile of the PBMCs from vaccinated individuals and found that most of the expressed mRNA genes were related to various signaling pathways of the innate and adaptive immune systems, and the immune functions were upregulated in comparison with the placebo group. Here, activation of the multiple signaling pathways involved in the immune response resulted in variations in hundreds of genes related to activation of innate immunity at day 7 after immunization regardless of the immunization schedule; however, the cytokines that were found to have elevated levels in COVID-19 patients had only mild variations and were at levels similar to those in the placebo control group, which corresponded with those detected in the serum. The activation of genes related to T cells, B cells, DCs and mononuclear cells/macrophages with varying dynamics is evidence of the immune response elicited by the vaccine. All the data obtained in this trial support the safety and immunogenicity of this inactivated vaccine and are encouraging with regard to further studies of its efficacy in the future.

A limitation of this study is the lack of protective efficacy analysis and data from a parallel contrasting analysis of the transcriptome of PBMCs from individuals immunized with the vaccine and of those from COVID-19 patients, which was due to a lack of samples.

## Data Availability

All data were available.The raw microarray data were submitted to the National Genomics Data Center (NGDC) and are available (PRJCA003531).

## Acknowledgments

We appreciate the contributions of all investigators at West China Second Hospital of Sichuan University who worked on the trial. This work was supported by the National Key R&D Program of China (2020YFC0849700), the Program of Chinese Academy of Medicine Science and the Major Science and Technology Special Projects of Yunnan Province.

## Author contributions

Conceived of and designed the experiments: QL, CL, YC, and LL. Performed the experiments: JP, QY, ZY, YZ, XL, DLi, ZZ, RL, HC, TM, HZ, SF, ZX, LW, ZH, YL, SF, JW, LZ, JL, HZheng, PC, GJ, LG, MX, HY, YG, XX, LC, LY, ZC, and DD. Analyzed the data: QL, CL, YC, LL, JP, YZ, DLi and ZZ. Contributed reagents/materials/analysis tools: QY, JR, JZ, CH, HZhao, YL, KM, DLiu, SL, XW and SY. Wrote the paper: QL, YC, YZ, DLi and ZZ. The manuscript was drafted by QL.

