## Supplemental Figure 1, 2 and Table 1 for "An in-depth investigation of the safety and immunogenicity of an inactivated SARS-CoV-2 vaccine"

***Supplemental appendix***

TABLE OF CONTENTS

|  | Page |
| --- | --- |
| **Methods** | 2-9 |
| Neutralizing antibody test | 2 |
| ELISAs | 2 |
| ELISpot assay | 2-3 |
| Isolation of immune cell populations | 3 |
| Cytokine detection | 3 |
| Transcriptome assay | 3-4 |
| qRT-PCR | 4 |
| ADE detection | 5 |
| Ethic | **6-9** |
| **Supplemental Figures** | **10-11** |
| Fig. S1 T cell populations in peripheral blood from individuals immunized with the inactivated vaccine | 10 |
| Fig. S2 The D614G mutation in the Spike protein of SARS-CoV-2 | 11 |
| **Supplemental Tables** | **12** |
| Table S1 Identification of the IgG antibody subtype against viral antigens elicited in immunized human individuals by the SARS-Cov-2 vaccine. | 12 |

**Methods**

***Neutralizing antibody test***

Heat-inactivated serum was diluted and coincubated with live virus (100 lgCCID_50_/well) for 2 h at 37°C, followed by the addition of Vero cells (10^5^/mL), and the mixture was incubated at 37°C in 5% CO_2_ for 7 days. The CPEs were observed and assessed to determine the neutralizing antibody titer of the serum. The GMTs of neutralizing antibodies were measured. Antibody titers ≥4 were considered positive. Seroconversion was defined as seropositivity after immunization in previously seronegative subjects.

***ELISAs***

S/N protein (Sanyou Biopharmaceuticals Co., Ltd., Shanghai, China) and purified viral antigen were used to coat 96-well ELISA plates (Corning, NY, USA) at a concentration of 5 μg/well and incubated at 4°C overnight. Then, the plates blocked with 5% BSA were incubated with serum samples and visualized with an HRP-conjugated antibody (Abcam, MA, USA) and TMB substrate (Solarbio, Beijing, China) as described in a previous report ^1^. The absorbance of each well at 450 nm was measured using an ELISA plate reader (Gene Company, Beijing, China). The antibody serum samples (which were diluted to 1:400) with an OD value (cutoff value) at least 2.1-fold higher than the negative control value were defined as positive. The endpoint titers (ETs) were defined as the highest serum dilution with a positive OD value. The geometric mean endpoint titer (GMET) was calculated as the geometric mean of the ETs of the positive serum samples in the same group. As for neutralizing antibodies, seroconversion was defined as seropositivity after immunization in previously seronegative subjects.

***ELISpot assay***

PBMCs of 50% of the subjects in the immunization or placebo groups were isolated from the blood via lymphocyte isolation (Ficoll-Paque PREMIUM; GE Healthcare, Piscataway, NJ, USA) and plated in duplicate wells. The stimulators of purified virion, recombinant S protein and recombinant N protein (Sanyou Biopharmaceuticals Co., Ltd.), were added to the wells. The positive control was phytohemagglutinin (PHA). The plate was incubated at 37°C for 24 h, the cells were removed, and the spots were developed. The colored spots were counted with an ELISPOT reader (CTL, Shaker Heights, OH, USA).

***Isolation of immune cell populations***

PBMCs were isolated via lymphocyte isolation (Ficoll-Paque PREMIUM; GE Healthcare). Anti-CD3, anti-CD20 and anti-CD16 antibodies were added to the PBMCs. The mixtures were incubated at room temperature (RT) for 30 min in the dark. Reagents for red blood cell lysis (BD) and membrane permeabilization (BD) were added in sequence. After two washes with PBS, the cells were resuspended in PBS and detected using a flow cytometer (BD, USA). T cells (CD3^+^), T helper cells (CD3^+^/CD4^+^) and CD8 (+) T cells (CD3^+^/CD8^+^) were evaluated. Furthermore, the T cells were categorized as T helper (Th) 1 and Th2 cells with anti-CD4, anti-IL-4 and anti-IFN-γ antibodies. The immune cell percentages were detected using a flow cytometer (BD). Th1 cells (CD4^+^/IFN-γ^+^) and Th2 cells (CD4^+^/IL-4^+^) were assessed.

***Cytokine detection***

The levels of 48 cytokines in the serum of the subjects were detected with a Th1/Th2 Cytokine Kit (BD) according to the manufacturer’s protocol. Briefly, serum was added to a tube containing detection beads. Then, the PE detection reagent was added, and the mixtures were incubated at RT for 2 h in the dark. After washing, the beads were resuspended in wash buffer and detected using a flow cytometer (BD). The levels of the cytokines were calculated according to a standard curve.

***Transcriptome assay***

Briefly, total RNA was extracted from PBMCs using an RNeasy Mini Kit (QIAGEN, GmBH, Germany). The RNA was checked for quality and quantified. Double- and single-stranded DNA and ribosomal RNA (rRNA) were removed. Magnetic beads were then used to purify and recover the reaction products. Subsequently, sequencing libraries were generated following the manufacturer’s recommendations. In brief, the reaction products were heated, denatured and circularized using a splint oligo sequence. Then, RNA-seq libraries were sequenced on an Illumina HiSeq X TEN platform (2×150-bp paired-end reads).

The read pairs were filtered using software to remove those with low-quality bases (Phred quality<5) or less than 10% uncertain bases. RNA-seq reads were aligned to the human genome

(http://ftp.ensembl.org/pub/release-84/fasta/homo_sapiens/dna/) using Bowtie V2.0.6. The raw count was used by DESeq2 to quantify the gene expression level. Cuffdiff was used to detect differentially expressed genes between the vaccine and control samples. Significant genes were identified at P<0.05.

***qRT-PCR***

RNA was extracted from samples using TRIzol reagent (Invitrogen Tiangen Biotech, China). Real-time RT-PCR assays were performed using a One-Step PrimeScript RT-PCR Kit (Takara, Shuzo, Japan) and a Real-Time PCR System (Bio-Rad, USA). The primers and probes of SARS-CoV-2 were used to measure mRNA levels: for N, forward: 5’-GGGGAACTTCTCCTGCTAGAAT-3’, reverse: 5’-CAGACATTTTGCTCTCAAGCTG-3’, and probe: 5’-TTGCTGCTGCTTGACAGATT-3’; and for ORF 1ab, forward: 5’-CCCTGTGGGTTTTACACTTAA-3’, reverse: 5’-ACGATTGTGCATCAGCTGA-3’, and probe: 5’-CCGTCTGCGGTATGTGGAAAGGTTATGG-3’. The primers and probes targeting the 3′ untranslated region (UTR) of DENV serotypes 1–3 were used according to previously described methods ^2^. An RT-PCR assay with universal single-probe for thediagnosis of dengue virus infections was performed as in the reference ^2^. The RT-PCR program was as follows: 42°C for 5 min, 95°C for 10 seconds, and 40 cycles of 95°C for 5 seconds.

***ADE detection***

Cells were infected with SARS-CoV-2 in the presence of either antiserum (which had a neutralizing titer of 1:32 and was diluted from 1:32 to 1:4096) from patients diagnosed with COVID-19 or from humans immunized with an inactivated vaccine or monoclonal antibodies (mAbs) against the SARS-CoV-2 proteins S and N. Supernatants and cell lysates were collected at different time points, and the viral load was determined by quantitative RT-PCR (qRT-PCR).

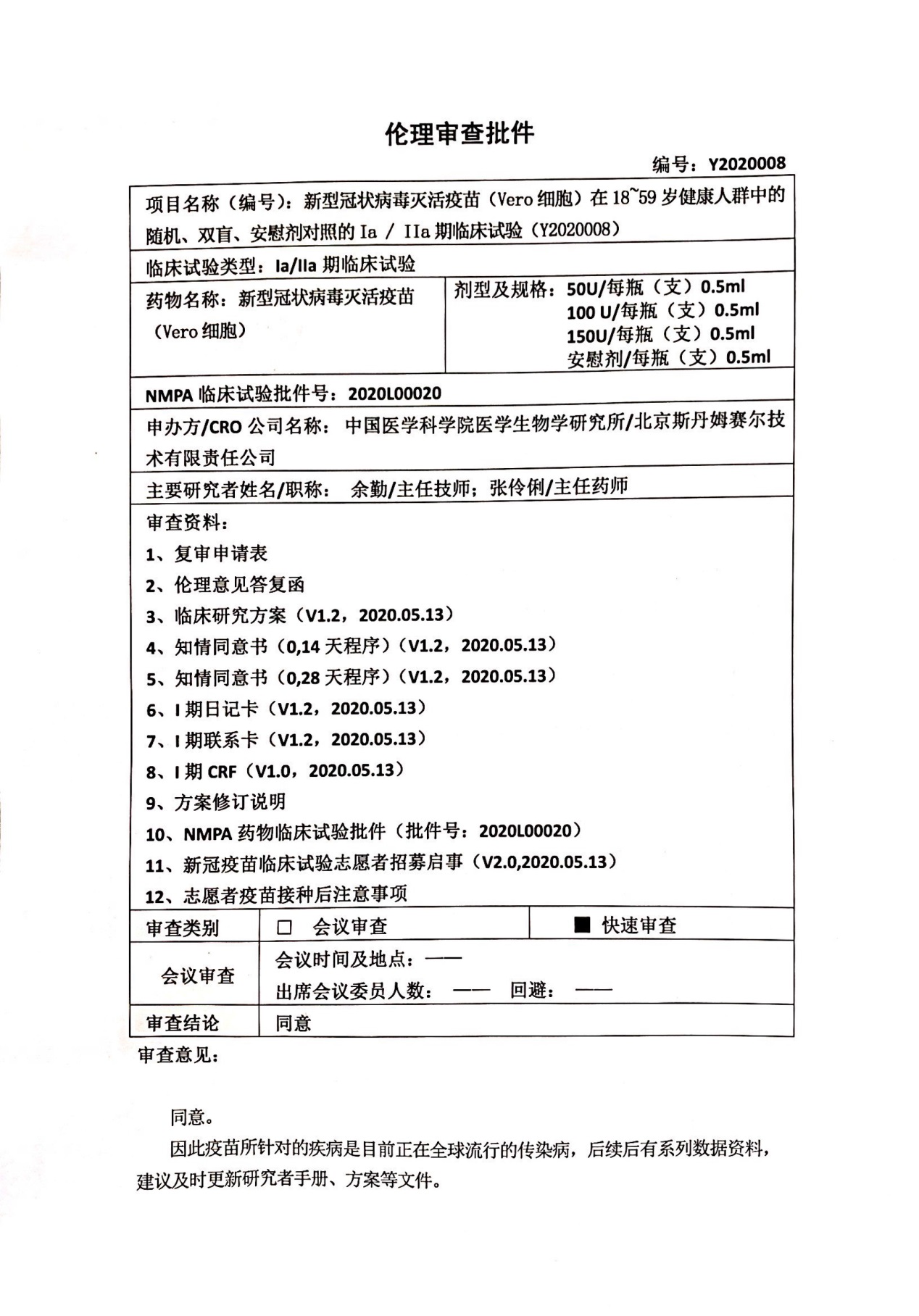

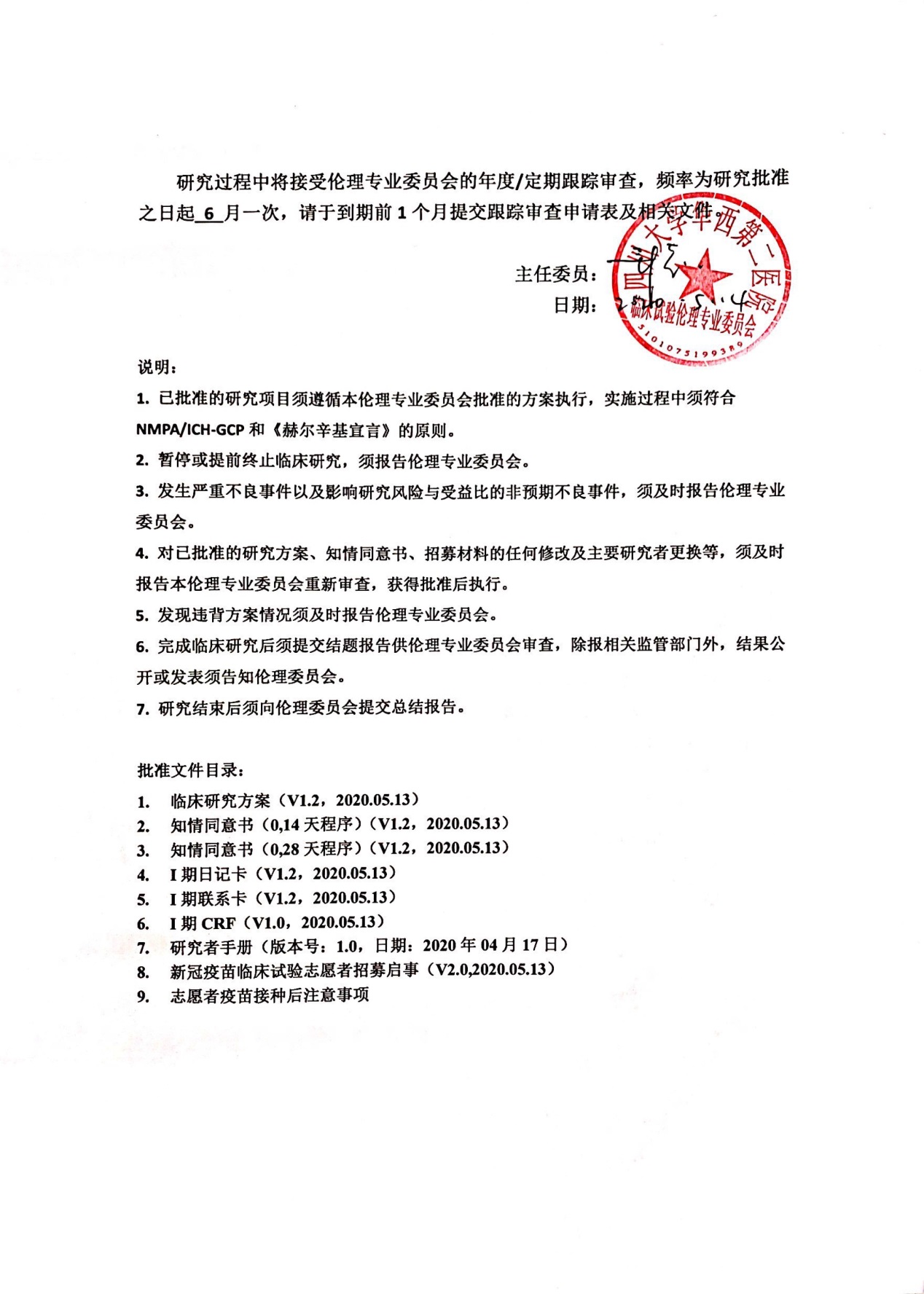

**Ethic Approval**

Approval No. Y2020008

Trial Name: A Randomized, Double-Blinded and Placebo-Controlled Phase Ia Trial of SARS-CoV-2 Vaccine, Inactivated (Vero Cell) in Healthy Population Aged 18-59 Years Old (Y2020008)

Trial type: Ia/IIa clinical trials

Drug name: SARS-CoV-2 Vaccine, Inactivated (Vero Cell)

Specification: 50 U/vial, 0.5 ml

100 U/vial, 0.5 ml

150 U/vial, 0.5 ml

placebo/vial, 0.5 ml

NMP approval No. 2020L00020

Sponsor/CRO: Institute of Medical Biology Chinese Academy of Medical Sciences/ Stemexcel Technology CO. LTD, Beijing

PI Name/Title: Qin Yu/Chief physician; Lingli Zhang/Chief physician

Reviewed documents:

1. Re-application form
2. Reply to the comments of the ethic committee
3. Study protocol (V1.2, 2020.05.13)
4. ICF (0, 14 procedure) (V1.2, 2020.05.13)
5. ICF (0, 28 procedure) (V1.2, 2020.05.13)
6. Diary card for Ia (V1.2, 2020.05.13)
7. Contact card for Ia (V1.2, 2020.05.13)
8. CRF for Ia (V1.2, 2020.05.13)
9. Explanatory notes to the protocol revision
10. NMP approval (No. 2020L00020)
11. SARS-CoV-2 vaccine trial recruitment notice (V2.0,2020.05.13)
12. Points for attention after receiving the vaccine by volunteers

Review type: quick review

Conclusion: approved

Review opinion: Approve to conduct the trial. Because this vaccine is targeted to control the COVID-19 pandemics in the global world, a number of subsequent research information shall be provided. It is suggested to update the investigator’s brochure and protocol etc. documents.

The total study implementation shall be followed up and reviewed based upon the annual/periodic requirement with a frequency of 6 month per time since the approval day. Please submit the follow-up review application form and other documents 1 month before the deadline.

Chairman (Signature)

Date: May 14, 2020

Notes:

1. The approved trial should be conducted in accordance with the protocol approved by our ethic committee, and with the principles as laid down in NMPA/ICH-GCP and Helsinki Declaration.
2. Suspend or terminate the trial should be reported to the ethic committee.
3. The occurrence of SAEs and SUSAR should be reported to the ethic committee.
4. Any revision to the approved protocol, ICF, recruitment notice and replacement of PI etc. should be reported to the ethic committee for review again, and can only be implemented after approval again.
5. Violation to the protocol should be timely reported to the ethic committee.
6. The summary report should be submitted to the ethic committee for review after study completion. The results publicly announced or published should be reported to the ethic committee in addition to the regulatory authorities.
7. The clinical study report should be submitted to the ethic committee after study completion.

Approved document lists:

1. Study protocol (V1.2, 2020.05.13)
2. ICF (0, 14 procedure) (V1.2, 2020.05.13)
3. ICF (0, 28 procedure) (V1.2, 2020.05.13)
4. Diary card for Ia (V1.2, 2020.05.13)
5. Contact card for Ia (V1.2, 2020.05.13)
6. CRF for Ia (V1.2, 2020.05.13)
7. Investigator’s brochure (V 1.0, 2020.04.17)
8. SARS-CoV-2 vaccine trial recruitment notice (V2.0,2020.05.13)
9. Points for attention after receiving the vaccine by volunteers

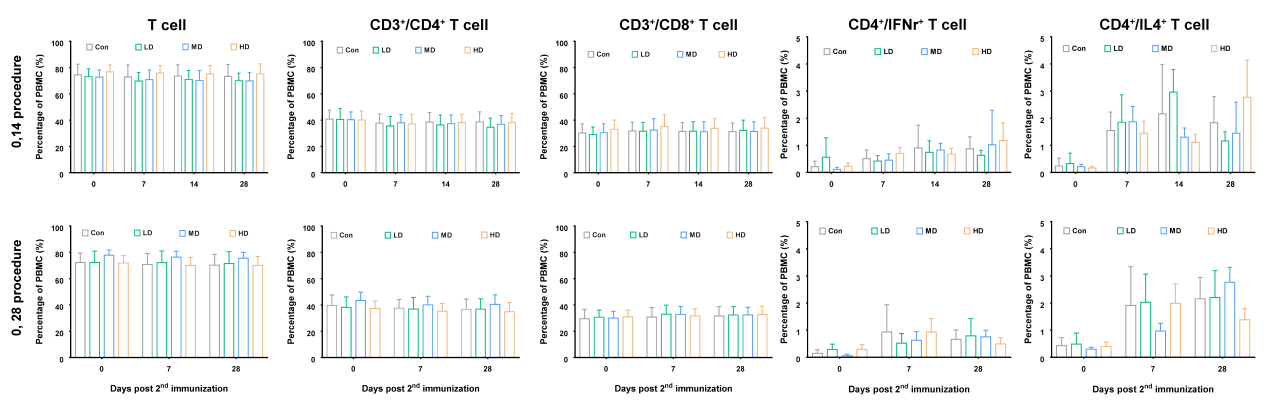

Fig. S1 T cell populations in peripheral blood from individuals immunized with the inactivated vaccine

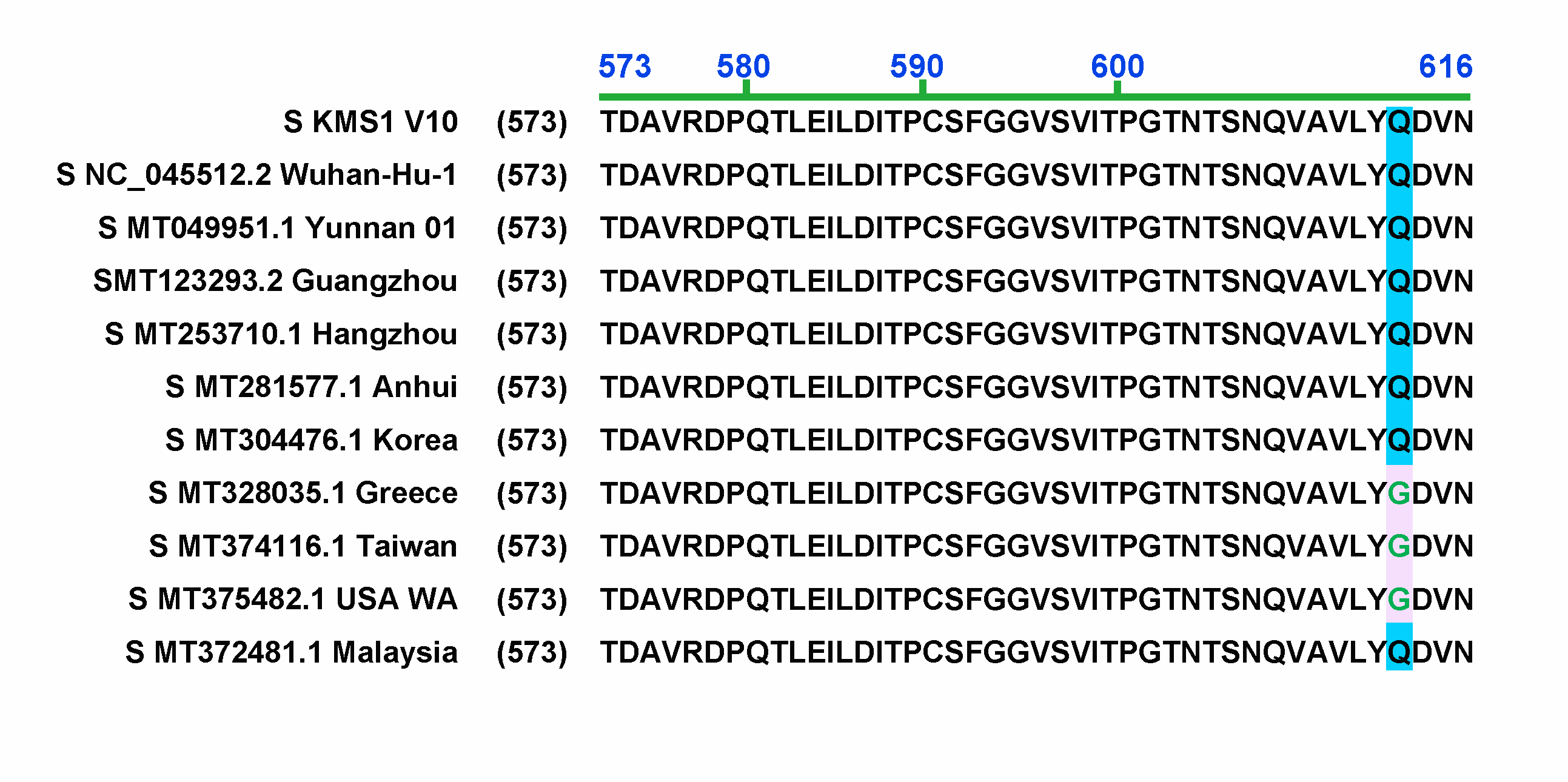

Fig. S2 The D614G mutation in the Spike protein of SARS-CoV-2

All nucleic acid sequences were derived from GenBank in NCBI.

**Table S1 Identification of the IgG antibody subtype against viral antigens elicited in immunized human individuals by the SARS-Cov-2 vaccine.**

|  |  | IgG1 | IgG2 | IgG3 | IgG4 |
| --- | --- | --- | --- | --- | --- |
| convalescent serum of patients | | | | | |
| S antigen |  | + | - | - | - |
| N antigen |  | + | - | - | - |
| Total antigen |  | + | - | - | - |
| immune serum from vaccinated individuals | | | | | |
| S antigen |  | - | - | - | - |
| N antigen |  | + | - | - | - |
| Total antigen |  | + | - | - | - |

Note: +, identified the antigen of SARS-CoV-2 (value of OD_450_ > cutoff value);

-, do not identified the antigen of SARS-CoV-2 (value of OD_450_ < cutoff value)
